# Improving communities based on the voices of children and young people in Wales: Findings from RPlace

**DOI:** 10.1101/2025.05.02.25326861

**Authors:** Levi Hughes, Michaela James, Marianne Mannello, Sinead Brophy

## Abstract

**Background:** The opportunity for children and young people (CYP) to play, socialise, explore and contribute within their community is fundamental to their wellbeing. CYP’s involvement in community development generates enthusiasm, motivation and a sense of belonging and significant national and international work is being undertaken to enhance child and youth participation. This paper explores what factors are important to CYP when they are involved in community development.

**Methods:** RPlace is a co-produced website and mobile application by CYP for CYP. CYP rate (from 1 to 5 using the Place Standard Tool) and review their local communities across six categories, including safety, activities and play, meet friends, accessibility, green space and cleanliness. Early pilot data for RPlace collated 311 responses from CYP, aged 7-24, from 13 local authorities in Wales.

**Results:** The results were analysed using thematic analysis and four key themes were identified. These themes were; i) being active and playing in communities *(“feels like one big community”)*, ii) being able to socialise in communities *(“you can’t meet friends as it is not safe”),* iii) ease of access to community spaces *(“I’d like to have easier and cheaper access to outdoor places”)*, iv) green space and clean environments *(“we need more green space and clean it up a little bit)*.

**Conclusion:** Involving CYP in community development can improve communities for all as they aim to increase safety, inclusivity and active travel and decrease antisocial behaviour, litter and dangerous driving. CYP participation in community development can be supported through RPlace, providing regular consultation through an anonymous, self-report and informal platform.

## Introduction

Community development should be rooted in collective action for social justice, promoting fundamental human rights and the importance of equality across local areas (1). The involvement of all individuals in this process, regardless of age or background, is essential to create shared goals (1, 2, 3). For children and young people (CYP), participation in community development can increase self-esteem through an empowerment to take ownership of their local spaces (1, 3). It has been acknowledged, however, that despite its focus on inclusivity, community development can overlook the views of minority groups in society, such as CYP (4). This has been echoed by CYP themselves who recall having limited autonomy to express and develop their own perspectives, to play a full part in decision-making processes and to make links between their ideas and potential impact (4, 5).

Limited opportunities for CYP to participate in community development are suggested to be the result of outdated perspectives on childhood (6) and the challenges associated with implementing child and youth participation into practice (7, 8). For example, it is widely recognised that CYP have been viewed as passive members of the community (9,10) whose voices and lived experiences have not been valued by those who carry the responsibility of listening to or advocating for them (11). Significant progress has been made as Article 12 of the United Nations Convention on the Rights of the Child (UNCRC) signifies and influences how all CYP now have the right to express their views and to have their views taken seriously (12). Internationally, research has been conducted on the implementation of the UNCRC worldwide (13) and from a national perspective, Article 12 underpins the work of key stakeholders in Wales as CYP are now recognised as active citizens with an important contribution to make to their communities (14). As reflected in the education system, recent changes to the *Curriculum for Wales (CfW)* educates all learners as capable, competent and curious citizens and the Ministerial Review of Play emphasises the importance of spatial justice for CYP, including their access to and participation in public spaces (15, 16).

Alongside the improvements made for the active citizenship of CYP, research emphasises the importance of tools and resources to accelerate CYP’s participation in community development as currently, there is a lack of clarity regarding how it should be carried out (7, 8). To support this growing body of work, RPlace is a website and mobile application that was co-developed with CYP. The purpose of RPlace stemmed from the ACTIVE Project at Swansea University that worked with over 1,000 teenagers (aged 13-14-years-old) who were given vouchers to spend on activities of their own choosing (19, 20, 21). It was found that whilst young people want unstructured, fun and social activities in their communities, they often felt unwelcome or that local provision was not made with them in mind. From this study, the research team collaborated with CYP to gain their perspectives on how their voices can have greater reach and impact and to design a way in which they could be represented in local spaces. Thus, RPlace was co-developed as an anonymous, self-report platform that enables CYP to rate and review their communities. This process can support local authorities and key stakeholders in gaining a deeper understanding of the direct thoughts, feelings and preferences of CYP to better inform policy and local provision in communities. This is a particularly important concept regarding play, for example, as researchers argue that public spaces and playground design do not adequately cater for CYP’s age-related needs or diverse experiences (17, 18).

The purpose of this study is to explore what factors are important to CYP when they are involved in community development, as gathered by RPlace, and to provide key recommendations from first and second stage piloting in South Wales, UK.

## Methods

RPlace was established in 2021 by researchers at Swansea University and in partnership with Play Wales as a result of conversations with CYP. The rating and review process includes closed-ended questions based on six different categories; safety, activity/play, meet friends, accessibility, green space and cleanliness. There is also an open-ended section where users can discuss these categories in greater detail or share any other strengths or weaknesses to their chosen area.

RPlace has been through two rounds of piloting in South Wales, the first in January-September 2021 and the second in May-October 2024. Throughout these stages, CYP were asked for their feedback and changes were made to the mobile application and website based upon their suggestions. For example, changes in the wording of prompts to allow for improved accessibility. This allowed RPlace to be fully co-produced with the involvement of its users.

Once the responses are submitted, reports can be generated from the findings and provided to local authorities or other key stakeholders to inform practice and policy. This creates a feedback-loop in which CYP’s voices can be represented.

### Data Collection

There are two ways in which data has been collected through RPlace. Firstly, the mobile application was designed and developed for users to enter or find the name of the area (established by a previous user) in their communities that they would like to rate and review. Users are then required to rate and review each of the six categories individually. These categories use the Place Standard Tool (22) and can be ranked 1 to 5 (1=worst). On the RPlace mobile application, there is also a feature for users to upload an image of their area to support their feedback.

Secondly and because of the feedback gained from participants, the RPlace website, designed for mobile and desktop use, was launched in 2023. On this platform, users enter the name of their chosen area, rate their area based upon the six categories and complete an open-ended review. These categories use the Place Standard Tool (22) and can be ranked 1 to 5 (1=worst). The deprivation level of each area on RPlace was identified using the Welsh Index of Multiple Deprivation (WIMD) (23). WIMD is a measure of multiple deprivation, and the domains used relate to material and social aspects of deprivation, including income, safety, housing and education (23). WIMD enabled the identification of trends regarding how users from different levels of deprivation perceived their local communities. The RPlace website also asks users for their age, gender and ethnicity. This section is optional; however, it is useful to provide stakeholders with demographic information about the CYP situated within their communities.

RPlace can be used independently or through school, play work and youth work engagement. Initially, it was envisaged that CYP would find the mobile app through social media promotion and collaboration with community organisations. However, piloting showed that engagement with schools, youth workers and local authorities is a helpful approach to recruiting CYP. This enables adults to create a space for CYP to engage with RPlace, to be on-hand for navigation or literacy support and overall, to promote the benefits of advocating for their communities.

The research team instigated this collaboration with schools and youth clubs in South Wales through in-person visits. For in-person visits, all schools and youth clubs are contacted via a recruitment phone call. Upon initial consent from the gate keeper, a suitable date and time is decided for the researcher to visit CYP. During the visit, the researcher presents a 20-minute presentation and demonstration of RPlace, asking CYP for their input and feedback. All classes are given an information sheet at the end of the session and CYP are provided with an information sheet to take home. For schools and youth clubs outside of South Wales, the research team instigates contact via email. If they are interested, they are sent a video walk-through and demonstration of RPlace that can be shared with classes. All information provided to schools regarding RPlace can be uploaded onto platforms for parents and guardians such as Class Dojo.

### Ethical Approval

RPlace has received ethical approval from Swansea University (#8808). Confidential treatment of user data is a key feature of RPlace. Users are encouraged to use pseudonyms for their usernames and passwords and should not include any confidential or identifiable information within the images they choose to upload to the RPlace app. All responses are monitored by the research team and are removed if they do not meet the appropriate guidelines. Written consent is obtained via the website and app at the start of the review submission. Swansea University’s Medical School Ethics Borad approved this consent procedure.

### Analysis

The reviews on RPlace were analysed using thematic analysis. As defined by Braun and Clarke (2006) (24), thematic analysis is a method used to interpret individual or group views, lived experiences and values. This method was best suited to RPlace data, supporting the researchers to identify common themes, ideas and repeated patterns that inform stakeholders on the thoughts, feelings and preferences of CYP regarding community development. This analysis was conducted by LH with supervision from MJ.

## Results

RPlace has received a total of 311 responses. Of this number, 249 responses were collected via the mobile application and 62 responses were collected via the website. These responses were representative of 12 of the 22 local authorities in Wales. This included Rhondda Cynon Taff, Caerphilly, Newport, Bridgend, Blaenau Gwent, Monmouthshire, Swansea, Carmarthen, Neath Port Talbot, Wrexham, Denbighshire and Powys. 73% of the responses were from those who identified as male and 22% were from those who identified as female (5% preferred not to say). 85% of the respondents were 7-11 years old, 8% were 11-15 years old and 6% were older. 67% of participants fell into the most deprived areas in Wales and 33% fell into the least deprived areas in Wales (23). The option to record demographic information was only available on the RPlace website.

Four themes were identified from the data. These themes were:

1. Being Active and Playing in Communities
2. Being Able to Socialise in Communities
3. Ease of Access to Community Spaces
4. Green Space and Clean Environments

### Theme 1: Being Active and Playing in Communities

This theme was the largest to emerge from the analysis and contained smaller sub themes relating to i) further opportunities and ii) safety concerns. Quotes from theme 1 can be seen in Table 1.

**Table 1.** Quotes from Theme 1.

|  |
| --- |
| **Theme 1** |
| *An absolutely amazing place, lots of outdoor and indoor activities like archery, horse riding, rock climbing and muddy assault courses, very welcoming and staff are really polite and helpful. A very lovely place to stay near the beach while having a country holiday as they have cottages in the centre also do water sports like paddle boarding. Lots of well looked after animals like horses, pigs and goats with lots of local wild animals like wild sheep and wild horses I would recommend Clyne farm as a family holiday, and place to meet with friends to do activities.* |
| *New playground, bowls, exercise equipment and football pitch.* |
| *I'd like more parks in general but I'd like bigger parks for older children to fit* |
| *More parks with fun things like monkey bars, skate parks or dog parks* |
| *I am really into sports so would like more pitches e.g. football and rugby pitches* |
| *This playground is in need of repair and redevelopment. The zip-line has been 'out of order' for as long as I can remember. The basket swing has been removed and many of the wooden trim trail parts have rotted - making the playground less playful for C&YP. Families from all across NPT come to visit Aberavon beach so it is disappointing that one of our most popular playgrounds is in such a sorry state.* |
| *There is no equipment accessible for a person in a wheelchair or any disabled person.* |
| *More clubs* |
| *A swimming center so be more active* |
| *Girls football, rugby, tennis teams and clubs* |
| *have free clubs and more places for us to play* |
| *Sometimes it feels dangerous in some streets* |
| *Stop people drinking at local parks* |
| *Safety is great except from 5:30 to 9:00 which is when the teenagers are coming out to meet up.* |
| *More zebra crossings* |
| *Need more traffic lights* |

Users spoke highly of the opportunities for activity and play that were available in their communities. One user praised the refurbishment of several playgrounds that have been funded by local councils and another user shared the success of activities in their area, such as archery, horse riding, rock climbing and the opportunity to see wild animals. On the other hand, an overwhelming majority of users discussed how they would like more opportunities to be active and play as they felt *“limited in what they can do”.* These responses included requests for additional parks, bigger parks and equipment that was more inviting for play, such as climbing frames, monkey bars, slides, swings, skateparks and football fields with goal posts. This was an important recommendation from one user in particular who felt *“disappointed”* that their local park and popular tourist attraction was in *“such a sorry state due to damaged, broken or rotten equipment”.* Several users also highlighted the little equipment that is available for CYP with disabilities, noting how *“there is no equipment accessible for a person in a wheelchair or any disabled person”*.

Alongside play and activity-based equipment, users discussed how they would like *“more opportunities to go to clubs”* within their local areas. It was especially important to users that these clubs were *“accessible to walk to”* and enabled *“outdoor play”* and the use of *“sports equipment outside of school”.* Examples included tennis clubs, football clubs for girls, running and walking clubs and swimming lessons. One user discussed how this would help CYP to *“spend less time on computers”* and to feel less *“lazy”.* As identified by WIMD (23), these responses were provided by users from areas of high and low deprivation.

Some safety responses referred to helpful ‘park keepers’ and how park gates were locked at night. A small number of users also reported feeling safe when walking alone as their local area *“feels like one big community”*. These positive safety responses were provided by users from areas of low deprivation in Wales. There was also a large proportion of negative safety responses regarding antisocial behaviour in communities and whilst users from areas of low deprivation reported concerns of bullying, users from areas of high deprivation reported *“knife crime and teenagers threatening”*, *“benches and equipment being set on fire and burnt down”, “people smoking, vaping, drinking and taking drugs”* and how *“you can’t meet friends as it is not safe”.* One user recalled how young people try to *“force”* children to participate in such activities and another stated how they would like to play outside of their home *“without getting hurt”*.

Other safety responses advised how communities would benefit from additional traffic lights, zebra crossings, speed cameras, streetlights, bike paths and walking paths. Such implementation would reduce users’ concerns regarding an increase in dangerous driving. Most of these recommendations were provided by users from areas of high deprivation.

### Theme 2: Being Able to Socialise in Local Communities

For some users, being able to use their communities encompassed having the time, space and freedom to socialise with their friends outside of the formality of being active. This involved having *“a community centre to meet friends outside of school”, “more benches”* and *“a sheltered place to hang out that has comfortable seats and lighting”.* These findings highlight how opportunities for informal play and socialisation are restricted for CYP due to limited resources. Quotes from theme 2 can be seen in Table 2.

**Table 2.** Quotes from Theme 2.

| Theme 2 |
| --- |
| *More free space* |
| *I would make a shop near where I live so I could have a chance to go to it without any supervision. so I can have some independence.* |
| *would like to have a community centre to meet up with friends* |
| *You can't meet friends as much as it is not safe.* |

### Theme 3: Green and Clean Spaces

Green space and clean spaces were two features that CYP people wanted and needed from their communities. The amount of green space that was available for activity and play varied for users. Some users discussed how they had limited space to *“run around in”* whereas others enjoyed playing in the *“big wide-open spaces”*, going on nature walks and visiting Welsh history landmarks. These responses were provided by users from high and low areas of deprivation. Few users also had access to blue space, including lakes and rivers. Two key recommendations regarding green space were for CYP to have access to *“larger garden areas for play”*, *“public fields”* and for *“more grass”* to prevent them from injuring themselves when playing on the equipment in parks. Quotes from theme 3 can be seen in Table 3.

**Table 3.**
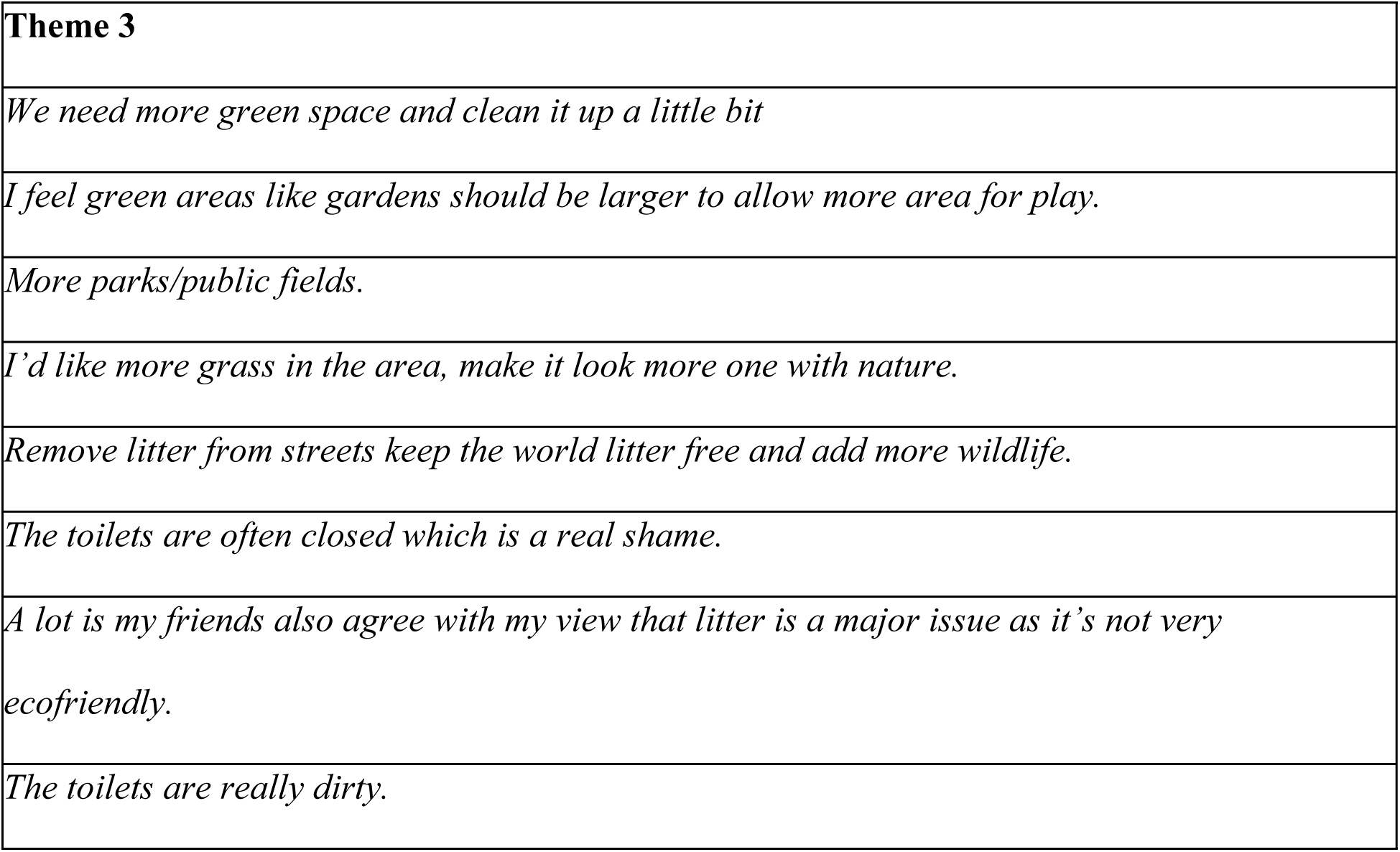
Quotes from Theme 3.

Nearly all users discussed the high volume of litter that is present within their communities and how spaces would benefit from more bins. Users also discussed the impact of vandalism, dog mess and rats on the appearance of their local areas. It was also discussed how park toilets are either *“always locked”* or *“dirty”* and *“never clean”.* Whilst users from all areas discussed litter, users from areas of low deprivation were more likely to provide recommendations or next steps on how to improve this concern. For example, one users discussed how they *“could do litter picking”* and another stated that *“there is a lot of talk about climate change and I feel to improve my area, we could do a lot more even locally like recycling, using local goods, even walking places rather than using cars, buses and trains, there is so much even a small community can do that would help improve the global warming crisis”*.

### Theme 4: Ease of Access to Community Spaces

For some users, it was important that a number of clubs and activities were made free of charge, *“have free clubs and more places for us to play”, “I’d like the price of things to be lower as things now a days have become quite expensive”.* These responses were recorded by users who live in one of the most deprived areas in Wales. For other users, it was important that clubs, activities, parks and leisure centres were within walking distance to their homes. This would reduce their concerns regarding the financial impact of petrol prices and public transport fees. For example, one user noted how *“transport nowadays is very expensive even travelling by car can be because of the expensive petrol prices, I’d like to have easier and cheaper access to outdoor places”.* Further quotes from theme 4 can be found in Table 4.

**Table 4.** Quotes from Theme 4.

| Theme 4 |
| --- |
| *I’d like the price of things to be lower as things now a days have become quite expensive.* |
| *Have free clubs and more places for us to play* |
| *Games that are free like frisbee to be more available.* |
| *Transport nowadays is very expensive even travelling by car can be because of the expensive petrol prices, I’d like to have easier and cheaper access to outdoor places.* |
| *I would change the local area by making everything free.* |

This theme emphasises the need for safer, more accessible and well-maintained local spaces for play, activity and socialisation. The concerns regarding safety and anti-social behaviour, limited opportunities, inadequate equipment and the lack of green space and clean environments reflect CYP’s desire to improve their local area.

## Discussion

This study explored the direct views of CYP regarding community development. The findings demonstrate the importance of providing CYP with a platform to share their valuable contributions and furthermore, their potential to positively impact provision, policy and community development. By listening to young people, communities can better address their needs for inclusive, affordable activities and safe, welcoming environments, ensuring that all youth, regardless of background, have the chance to use their local spaces equally.

### Improving Opportunities to Play and Be Active in Communities

Playing and being active within their communities was significant for CYP. Such opportunities were impeded for RPlace users, however, as they noted feeling unsafe at local parks. This finding aligns with wider research that states how crime rates and CYP’s low levels of safety within communities contribute towards the decline in outdoor play (3). To increase CYP’s play and activity within their local areas, it is recommended that local authorities, councils and key organisations target the social connectedness of communities and the social bonds between neighbours (3). The Welsh Government promotes Play Streets as a potential initiative to achieve this (24). Play Streets permit temporary street closures for community and sporting events, thereby increasing the opportunities that CYP have to play outside and make friends (24). Where Play Streets have taken place in Wales, residents and organisations such as Play Wales shared their success and discussed how they can reduce anxieties, create a greater sense of cohesion and develop more collaborative communities (25, 26). Play Streets also have the potential to reduce the safety concerns of RPlace users regarding the increase in dangerous driving as they enable CYP to play outside and away from traffic (25).

To further improve the opportunities that CYP can play and be active within their communities, RPlace has gained insight into what they believe meaningful play is. For example, many users noted their enthusiasm for more exciting equipment in parks and playgrounds, including the use of equipment that has not been damaged or broken. These responses are supported by researchers who argue that litigation fears restrict them to boring and unappealing post and platform-based structures (17). International responses to this widespread concern include the introduction of innovative parks and playgrounds that encourage CYP to have more unique experiences (17).

These spaces are designed for all age-groups and consist of loose and moveable equipment, natural and planted areas that encourage play and open-ended structures that do not direct play sequences (17). The implementation of more engaging equipment is a key recommendation for CYP as play enables curiosity and exploration, abstract thinking and the opportunity to build resilience and release emotions (27, 28). An increase of naturalised and planted areas for play would be useful as the results of RPlace and wider research demonstrates that outdoor play, particularly that within green and blue space, is the optimal environment for CYP to experiment with movement and enhance their holistic well-being (28, 29). Outdoor play is also more widely associated with community play, therefore enabling CYP to increase their participation in community development, to explore and learn life lessons within their local areas and to develop positive relationships with adults (3).

Amongst the responses regarding play-based equipment in communities, several users discussed the lack of equipment that is available to disabled CYP. This is a widespread issue as researchers emphasise the ongoing exclusion of these individuals (18). Despite the global commitment to inclusion, this experience means that CYP with disabilities spend less time outdoors, have reduced opportunities for self-regulation and in turn, are more likely to be associated with the rise in childhood and adolescent psychological and behavioural disorders (30, 31, 32). International observations suggest that stakeholders often have insufficient knowledge in this area due to a lack of guidance on how to create accessible and usable playgrounds for all CYP (33). As a result of this information, it is argued that extra work is required to support CYP with disabilities in exercising their right to play and to express their opinions in regard to community development (18). As demonstrated in the results, RPlace is a useful tool to support this movement and to hear the direct views of CYP with disabilities and the views of their friends and families.

### Being Able to Socialise in Communities

Being able to socialise in communities offers a wide range of benefits for CYP. Social interactions in public outdoor spaces are formative to psychological well-being and they provide CYP with the opportunity to explore their independence and to construct personal and social identities (3, 34). This was exemplified in the findings where RPlace users highlighted the importance of socialising outside of the formalities of play and activity.

Whilst hanging out with friends enables all CYP to form deeper connections, researchers acknowledge the impact of gender inequalities on this process (34). For example, provision of outside space is often occupied by football pitches and empty fields that have the potential to be boy-dominated (34). It is important to note here that whilst many girls engage in and enjoy these activities, design movements now aim to better support their unique needs through smaller spaces, improved lighting, clear sightlines and areas for social seating (3, 34, 35). Similar arguments exist in relation to socioeconomic differences and how this influences CYP’s ability to hang out and socialise within their communities (36). For example, it has been discussed how in comparison to their more affluent peers, CYP from deprivation often find difficulties in making friends and maintaining friendships (36, 37). Similarly, more deprived communities can restrict the independent mobility of CYP and increase sedentary behaviours through a lack of recreational, safe and green space (37).

### Green Space and Clean Spaces

It was identified in the findings how high volumes of litter can make communities unwelcoming for CYP. This was of high concern for users from more affluent areas who wanted to reduce litter and its impact on climate change through recycling and litter picks. This finding aligns with wider research that identified how CYP are more likely to utilise well maintained safe areas and play in areas where they have been involved in the maintenance process (38, 39, 40). There are also over two hundred litter picking hubs across Wales that provide free equipment for safe clean-ups of communities (41). This is an initiative that could be driven by schools, replicating successful projects such as Litter Pick and Play that aim to empower CYP to advocate for their right to clean spaces (40).

### Ease of Access to Community Spaces

A frequent response throughout the findings was the request for more parks, clubs and activities to be within walking distance of users’ homes. One user also discussed how they would like to walk to school to become healthier and fitter. These responses highlight the importance of active transportation for CYP in Wales. Active transportation refers to modes of travel that involve physical activity and it plays a key role in improving physical, mental and social well-being (42). As identified by researchers in Wales however, whilst 73% of CYP used active transportation to travel to places where they play, only 35% of children and 44% of young people used active transportation to travel to school (43). These statistics align with wider research that evidences the increase in car-centric cultures and passive transportation to and from education and leisure related destinations (44, 45).

Whilst it is not always feasible to introduce additional parks, clubs and activities across all areas of Wales in the short-term, stakeholders have discussed the potential impact of routes that encourage active travel. For example, the Welsh Government, through their discussion on spatial justice, reinforced the need for more well-designed and people orientated streets including the roll-out of safer and more attractive routes for walking, cycling, formal and informal play (15). It was further discussed by the Welsh Government how this roll-out would increase CYP’s independent mobility; their everyday freedoms to explore their natural world and to engage in outdoor play without adult supervision (15). This would be a positive movement as the independent mobility of CYP has decreased over recent years due to the large proportion of schools and leisure activities that are now situated outside of immediate communities (42, 44, 45). Researchers state that this is an undesirable trend as active travel enhances CYP’s sense of direction and is environmentally smart, reducing noise and air pollution and carbon emissions (42, 44, 45).

Future work could look to pilot the roll-out of improved active travel routes within an area where RPlace users requested additional opportunities to walk to education or leisure related destinations. Thus, the RPlace website and app could collect data on the thoughts and new experiences of these individuals, identifying whether it has enabled CYP to increase their participation in active transportation. This pilot would also be useful for users who expressed concerns regarding petrol costs and public transportation fees.

### Limitations

As discussed in the methods, RPlace has been through two rounds of piloting in South Wales. This process has enabled the research team to identify potential issues and make necessary adjustments. Three key issues were identified. Firstly, the option to record demographic information was only available on the RPlace website. Secondly, whilst the RPlace website and app enables users to rate their communities using the Place Standard Tool, these ratings were not included within the results of this paper as a large proportion of users misinterpreted the Place Standard scoring system. For example, positive reviews often received a rating of 1 (very bad). To avoid this situation in the future and to improve the reliability of data, RPlace, going forward, will use emojis (a happy face for good, a straight face for average and a sad face for bad). This will create an easier and more engaging experience for users. Thirdly, the RPlace data was only representative of white CYP in Wales. For future data collection purposes, researchers aim to target communities with a greater level of diversity.

## Conclusion

This study has explored what is important to CYP when they are involved in community development as identified by the RPlace app and website. RPlace has been implemented as an effective tool to accelerate CYP participation in community development and to support the growing body of work that promotes CYP as active citizens who have the right to be heard and taken seriously. The key message from this study is the importance of fully integrating CYP into community life, ensuring that these individuals are provided with a platform to act upon their desire to improve their local area. Given the findings, there are three key recommendations to enhance this experience:

1. To ensure that public policy in Wales is guided by the autonomy voices and input of CYP. This can be achieved through bi-annual council and local authority-led consultations with CYP and the use of evidence from RPlace. In doing so, the voices of CYP will be heard on decisions, policy reviews and policy implementations. This is an important consideration regarding funding as the RPlace results and wider research (3) indicate that play and leisure facilities are inefficient without the input of CYP during planning and development stages.
2. Introduce additional road safety measures. RPlace results identified that road safety concerns were one of the greatest barriers to CYP’s outdoor play and socialisation. It was discussed how this could be addressed through the introduction of additional traffic lights, zebra crossings, streetlights and paths near play areas.
3. To improve the maintenance of communities. This recommendation could be achieved through the active involvement of CYP in the maintenance of public spaces. This would be a low-cost initiative driven by schools and its relevance to the Health and Well-being and Science and Technology Areas of Learning and Experience (AoLE).

Alongside CYP, these recommendations have the potential to benefit the wider population. For example, CYP who have the time, space and freedom to play and socialise are more likely to act positively and wish to contribute to the development of their communities, thereby supporting current and future generations to become knowledgeable about their community and wider society, to learn life skills through their environment and to build positive social relationships with adults.

Overall, this study has demonstrated what is important to CYP when they are involved in community development. RPlace can increase child and youth participation in community development as it provides regular consultation with children and young people through an informal, anonymous and self-report platform.

## Data Availability

Data cannot be shared publicly because of the qualitative nature of the dataset and the potential for participant identification. Data are available from the Swansea University Institutional Data Access / Ethics Committee (contact via) for researchers who meet the criteria for access to confidential data.

## Acknowledgements

We would like to sincerely thank the children and young people who contributed their time, thoughts, and ideas through the RPlace platform. We also gratefully acknowledge the support of schools, youth groups, and local authorities across Wales who facilitated participation. Special thanks to Play Wales for their partnership and ongoing advocacy for children’s rights to play and participate in community life.

## Notes

### Competing Interest Statement

The authors have declared no competing interest.

### Funding Statement

Yes

### Author Declarations

RPlace has received ethical approval from Swansea University Medical School Ethics Board (#8808).

